# Toward Assessing Clinical Trial Publications for Reporting Transparency

**DOI:** 10.1101/2021.01.12.21249695

**Authors:** Halil Kilicoglu, Graciela Rosemblat, Linh Hoang, Sahil Wadhwa, Zeshan Peng, Mario Malički, Jodi Schneider, Gerben ter Riet

## Abstract

**Objective:** To annotate a corpus of randomized controlled trial (RCT) publications with the checklist items of CONSORT reporting guidelines and using the corpus to develop text mining methods for RCT appraisal.

**Methods:** We annotated a corpus of 50 RCT articles at the sentence level using 37 fine-grained CONSORT checklist items. A subset (31 articles) was double-annotated and adjudicated, while 19 were annotated by a single annotator and reconciled by another. We calculated inter-annotator agreement at the article and section level using MASI (Measuring Agreement on Set-Valued Items) and at the CONSORT item level using Krippendorff’s *α*. We experimented with two rule-based methods (phrase-based and section header-based) and two supervised learning approaches (support vector machine and BioBERT-based neural network classifiers), for recognizing 17 methodology-related items in the RCT Methods sections.

**Results:** We created CONSORT-TM consisting of 10,709 sentences, 4,845 (45%) of which were annotated with 5,246 labels. A median of 28 CONSORT items (out of possible 37) were annotated per article. Agreement was moderate at the article and section levels (average MASI: 0.60 and 0.64, respectively). Agreement varied considerably among individual checklist items (Krippendorff’s *α*= 0.06-0.96). The model based on BioBERT performed best overall for recognizing methodology-related items (micro-precision: 0.82, micro-recall: 0.63, micro-F1: 0.71). Combining models using majority vote and label aggregation further improved precision and recall, respectively.

**Conclusion:** Our annotated corpus, CONSORT-TM, contains more fine-grained information than earlier RCT corpora. Low frequency of some CONSORT items made it difficult to train effective text mining models to recognize them. For the items commonly reported, CONSORT-TM can serve as a testbed for text mining methods that assess RCT transparency, rigor, and reliability, and support methods for peer review and authoring assistance. Minor modifications to the annotation scheme and a larger corpus could facilitate improved text mining models. CONSORT-TM is publicly available at https://github.com/kilicogluh/CONSORT-TM.

**Graphical abstract:** 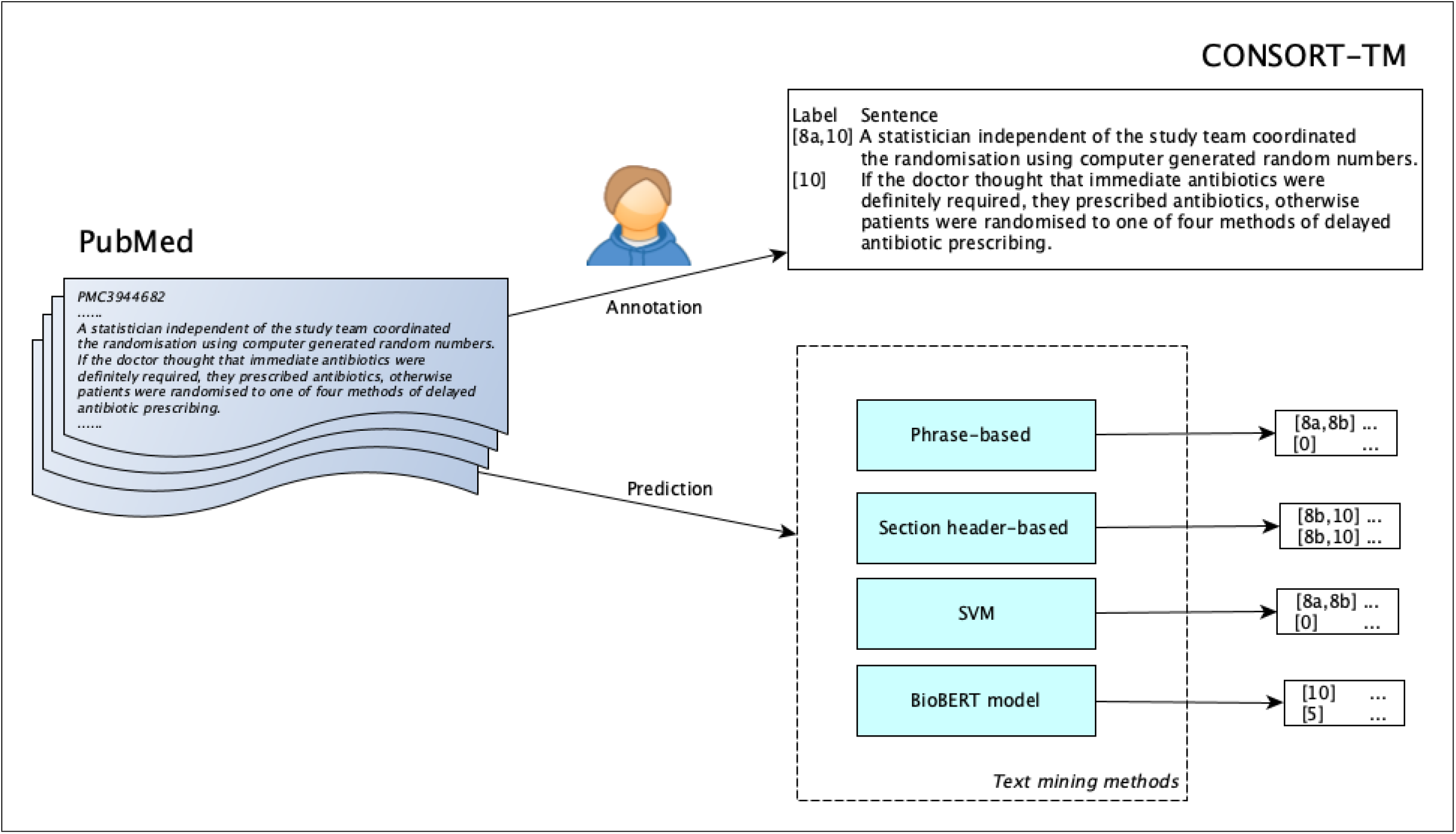

**Highlights:**

- We constructed a corpus of RCT publications annotated with CONSORT checklist items.
- We developed text mining methods to identify methodology-related check-list items.
- A BioBERT-based model performs best in recognizing adequately reported items.
- A phrase-based method performs best in recognizing infrequently reported items.
- The corpus and the text mining methods can be used to address reporting transparency.

## 1. Background

Rigor and reproducibility of scientific research has been widely questioned in recent years [1]. The biomedical research enterprise is at the center of these discussions and concerns have been raised about research waste in biomedicine [2]. To address these concerns, various interventions (e.g., grant application requirements [3], transparency and openness guidelines [4], data sharing principles [5], peer review of study protocols [6]) have been proposed.

Incomplete reporting and lack of transparency is a common problem in biomedical publications and can hinder efforts to replicate prior research findings because important methodological details may be missing [7, 8]. When key elements such as sample size estimation, randomization/blinding procedures or study limitations are not reported, it can also be difficult to assess the rigor and reliability of a study for evidence synthesis. While transparent reporting alone does not guarantee a rigorous and reliable study, it is an essential step toward assessing a study for such criteria [9]. Conversely, a well-executed but poorly reported study is unlikely to pass a thorough peer review process.

Reporting guidelines have been proposed to improve transparency and accuracy of reporting in publications resulting from biomedical studies. These include CONSORT for randomized controlled trials (RCTs) [10], ARRIVE for pre-clinical animal studies [11], and PRISMA for systematic reviews [12], all developed under the umbrella of the EQUATOR Network [13]. While they have been endorsed by many high-impact medical journals [14], adherence to reporting guidelines remains sub-optimal [15].

RCTs are considered a cornerstone of evidence-based medicine [16] and are placed high in the “evidence pyramid” [17]. To fully exploit their theoretical advantages, they need to be rigorously designed and conducted, and clearly and accurately reported. The CONSORT Statement for RCT reporting, developed in 1996 [10] and updated in 2010 [18, 19], consists of a 25-item checklist and a participant flow diagram. CONSORT has been extended over the years to RCT abstracts [20] as well as to specific types of RCT designs, such as cluster trials [21], and interventions, such as non-pharmacologic treatments [22]. CONSORT is the best known reporting guideline, endorsed by 585 biomedical journals^1^ and prominent editorial organizations. Prior research has shown that journal endorsement was correlated with completeness of reporting, while the reporting of key methodological details, such as allocation concealment, was lacking even in endorsing journals [23].

Studies assessing adherence to CONSORT rely on manual analysis of a relatively small number of publications [23]. A text mining tool that can locate statements corresponding to CONSORT checklist items could facilitate adher-ence assessment at a much larger scale. Such a tool would also have broader utility. For example, it could assist authors in ensuring completeness of their reports, journal editors in enforcing transparency requirements, and peer reviewers, systematic reviewers, and others in critically appraising RCTs for transparency and rigor criteria [24].

Most modern text mining methods need to be trained on large amounts of representative, labeled text to be successful. Developing such corpora based on the CONSORT guidelines is a challenging task, given the large number of checklist items, and the expertise required for identifying statements that correspond to these items in RCT publications. In this study, we aimed to create a corpus of RCT articles annotated with CONSORT checklist items at the sentence level and use it to train and evaluate text mining methods. Herein, we describe the annotation process, corpus statistics, and inter-annotator agreement for the corpus (named CONSORT-TM). We also present baseline experiments in classifying sentences from RCT articles with these items. We limit these experiments to Methods sections and methodology-specific CONSORT items, covering key methodological details most relevant to trial rigor and robustness.

## 2. Related Work

Text mining research on RCT articles has primarily concentrated on annotating and extracting study characteristics relevant for article screening for systematic reviews and evidence synthesis [25, 26].

Much attention has been paid to PICO elements (Population, Intervention, Comparator, and Outcome), used in evidence-based medicine [16] to capture the most salient aspects of clinical intervention studies [27–30]. PICO elements were annotated at the noun phrase level [27, 29] as well as the sentence level [27, 28]. Semi-automatic methods [28] and crowdsourcing [29] have also been used to generate training data. For example, EBM-NLP corpus [29] consists of 5000 abstracts, annotated at the text span level through crowdsourcing. In their corpus, some PICO elements are further subcategorized at more granular levels (e.g., Physical Health as a subcategory of Outcomes). Text mining methods used to extract PICO characteristics ran the gamut from early knowledge-based and traditional machine learning methods [27] to more recent semi-supervised [28] and neural network models, including Long Short-Term Memory (LSTM) and recurrent neural networks (RNN) [29, 30]. PICO variants, such as PIBOSO (B: background, S: study design, O: other), have also been considered. A corpus of 1000 abstracts annotated at the sentence level with PIBOSO elements has been published (PIBOSO-NICTA) [31], and various machine learning models (conditional random fields, neural network models) trained on this corpus have been reported [31–34].

Other research looked beyond PICO and its variants. Kiritchenko et al. [35] annotated 21 elements from 132 full-text articles, including eligibility criteria, sample size, and drug dosage. To automatically recognize these elements, they used a two-stage pipeline which consisted of machine learning-based sentence classification followed by phrase matching based on regular expressions. Hsu et al. [36] focused on information related to statistical analysis in full-text articles (hypothesis, statistical method, interpretation) and used rule-based methods to identify these elements in a dataset of 42 full-text articles (on non-small-cell lung carcinoma) and map them to a structured representation. Marshall et al. [37] identified risk-of-bias statements in RCT publications and categorized the studies as high or low risk with respect to risk categories, including sequence generation and allocation concealment. Their dataset was semi-automatically generated from the Cochrane Database of Systematic Reviews, and they used support vector machine (SVM) classifiers to jointly learn the risk-of-bias levels and the supporting statements in the article [37]. We have previously developed methods to automatically recognize limitation statements in clinical publications using a manually annotated corpus of 1257 sentences [38].

Annotation and extraction of key statements has also been considered for other publication types, such as pre-clinical animal studies and case reports [39–42]. One work that is particularly relevant to ours is SciScore [42], a tool which assesses life sciences articles for rigor and transparency, by extracting characteristics including subject’s sex, sample size calculation, institutional review board statements, antibodies and cell lines, and calculating a score based on them. They manually annotated several datasets to train named entity recognizers; however, these datasets are not publicly available, to the best of our knowledge.

Our work extends earlier research in several ways. Owing to our focus on CONSORT, we annotated a greater number of study characteristics, some of which have not been annotated in any publicly available dataset, to our knowledge. As a result, our dataset is more suitable for training text mining methods that address transparency more comprehensively. We also believe that methods trained on CONSORT-TM can be useful for a broader set of tasks than systematic review screening and authoring. In addition to traditional rule-based and supervised learning approaches, we also experimented with neural network classifiers based on contextualized language models, specifically BioBERT [43].

## 3. Materials and Methods

### Corpus Annotation

We manually annotated 50 articles from 11 journals with 37 fine-grained CONSORT checklist items (the complete checklist and the list of 11 journals are provided in Supplementary file 1)^2^. We used a modified version of Cochrane’s sensitivity and precision-maximizing query for RCTs as the search strategy^3^ to sample RCT articles for annotation. We limited the search to articles published in 2011 or later, since the most recent CONSORT statement was published in 2010. Our search resulted in 563 articles (search date: July 16, 2018). We randomly selected a convenience sample of 50 articles for annotation.

We downloaded PubMed Central (PMC) XML files of the articles and processed them with an in-house sentence splitter and section recognizer to identify the sections and the sentences in each section. In addition to the title, abstract, and the full text of the article, we also extracted its back matter, as some CONSORT items are often stated in this section (e.g., Funding (25)).

The annotation task consisted in labeling sentences of the articles with relevant CONSORT items. First, two authors (HK and GR) labeled sentences in one article using the CONSORT explanation and elaboration article [19] as their guide. These annotations were then discussed and adjudicated and were provided as an example annotated document to all annotators. They also formed the basis of the preliminary annotation guidelines. Next, six annotators double-annotated 30 articles independently (10 articles per annotator). The annotators are experts in meta-research (with medical degrees), clinical trial methodology, biomedical informatics, text mining, and linguistics, all well-versed in scholarly communication, specifically biomedical literature. The annotators were expected to read the guidelines and comment on them before beginning annotation. The guidelines were iteratively updated throughout the study, as needed (the guideline document is provided in Supplementary file 1). One annotator labeled a single article due to time constraints, their remaining articles were assigned to other annotators, while still ensuring double annotation of each article. Each of the 30 articles were then adjudicated by one of the two authors (HK or GR). Next, another 19 articles were annotated by a single author (GR), whose annotations were inspected and corrected by the first author (HK).

We used a custom, web-based annotation tool for labeling articles. The tool allowed the annotator to navigate the article using tabs (corresponding to article sections) and select labels for each sentence from a drop-down list. A link to the annotation guidelines was provided, as well as a link to the PDF of the article for better contextualization and access to tables and figures. The user was afforded the ability to check which items they had and had not annotated in the article (Item Check button). A screenshot of the annotation interface is shown in Figure 1. Annotation adjudication was performed using a modified version of the same tool.

**Figure 1:**
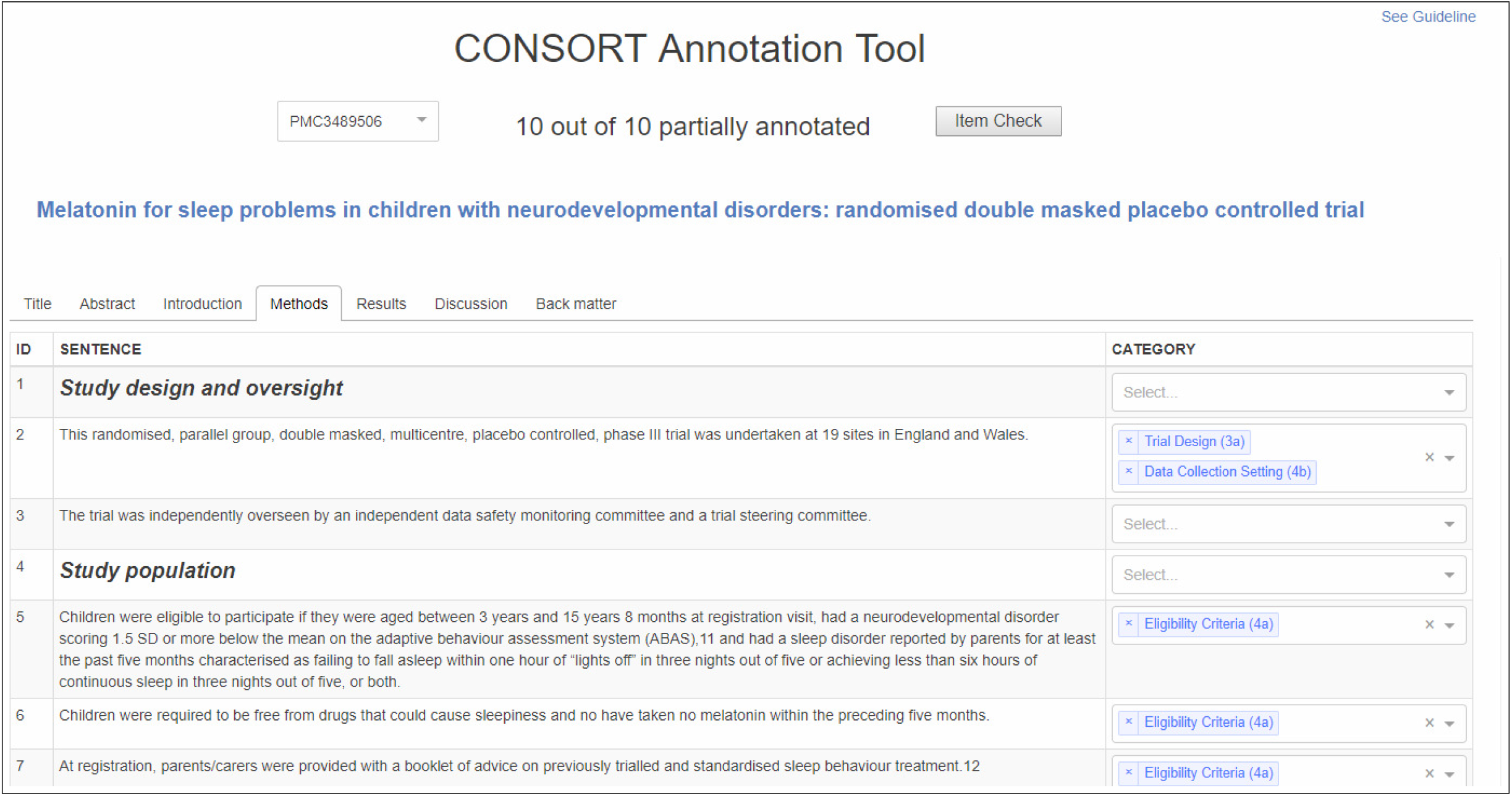
CONSORT-TM annotation interface

### Inter-annotator agreement

We calculated inter-annotator agreement for 30 double-annotated articles. Since each sentence could be labeled with multiple items, we chose MASI measure (Measuring Agreement on Set-Valued Items) [44] to calculate agreement. MASI is a distance metric for comparing two sets and incorporates the Jaccard index. It penalizes more the case in which two compared sets have disjoint elements than the case in which one set subsumes the other. We calculated pairwise inter-annotator agreement using MASI at the article and section levels. We also calculated Krippendorff’s *α* [45] for each CONSORT item. This measure was chosen since the annotation involved more than two annotators and the annotation data was incomplete (i.e., not all sentences were labeled by all annotators). We excluded CONSORT items annotated at the article level (1a and 1b) from agreement calculations. We also excluded some sentences (title, abstract, and section headers), which the annotators were instructed not to annotate, from these calculations.

### Text mining methods

We cast the problem of associating sentences with CONSORT items as a sentence-level, multi-label classification task. In this study, for text mining, we specifically focused on the Methods section of RCT articles and the 17 fine-grained items associated with this section in CONSORT guidelines (items 3a to 12b), as they are highly relevant for assessing rigor and reliability of a clinical trial.

### Rule-based methods

Our rule-based methods were based on two observations:

- When articles are organized by subsections, the subsection headers can provide clues about the items discussed in those sections (e.g., A subsection with the header *Participants* is likely to contain sentences discussing Eligibility Criteria (4a)).
- Certain words/phrases in a sentence are clues for particular CONSORT items (e.g., *block size* is likely to indicate a sentence on Sequence Generation (8a)).

To identify such patterns, we collected the list of all Methods subsection headers from a separate, larger set of RCT articles from PMC (about 18K articles with a clinicaltrials.gov registry number), ranked them by their frequency, and mapped the most frequent ones (or their relevant substrings) to CONSORT items. For rare checklist items (e.g., Changes to Outcomes), we also examined the long tail to identify at least a few relevant headers. An example mapping relates the string *concealment* to the CONSORT item Allocation Concealment (9). We assign this label to the sentences from the sections with the following headers: *Allocation concealment mechanism, Concealment of allocation, Concealment of group allocation to participants*. This method uses 48 mappings for 17 Methods-specific items. For phrase-based classification, we generated a set of predictive phrases, including *power to detect* for Sample Size Determination (7a) and *masked to treatment* for Blinding Procedure (11a), from the subsections with the headers corresponding to CONSORT items (a total of 232 phrase mappings). If a sentence contains phrases associated with a particular CONSORT item, we label the sentence with that item.

### Supervised machine learning

Two other methods were based on supervised machine learning. One approach involved a support vector machine (SVM) classifier that uses as features *tf-idf* representation of the sentence in addition to the enclosing subsection header. The section header was prepended to the sentence and included in *tfidf* calculation. We excluded common English words using the NLTK stopword list and used the LIBLINEAR SVM implementation in the `scikit-learn` package ^4^. C regularization parameter was set to 10 after a grid search. The classifier was embedded into a one-vs-rest classifier to enable prediction of multiple labels for each sentence.

The other approach involved a neural network classifier based on BioBERT [43], a variant of the BERT model [46] pre-trained on the biomedical literature. BERT is a bidirectional Transformer [47] trained on language modeling tasks over massive datasets in an unsupervised manner. The BERT encoder produces a vector of hidden states from text input, which can then be fine-tuned for a supervised task, such as sentence classification. BERT has been shown to yield state-of-the-art results on many NLP tasks in recent years, even with small datasets, motivating its use in this study. BioBERT was pre-trained on PubMed abstracts and PMC full-text articles in addition to original BERT training data. Sentence text and its subsection header were fed as input to the BioBERT encoder, whose output was then used to train the final sigmoid layer for multi-label classification. We used the `simpletransformers` package to implement multi-label text classification^5^. The following hyperparameters were used for model training and evaluation: batch size (4), learning rate (3e-5), number of epochs (30), optimizer (Adam), dropout (0.1).

## 4. Results

### Corpus annotation

The descriptive statistics of CONSORT-TM are given in Table 1. The corpus contains over 10K sentences, 45% of which were annotated (4845 sentences). The total number of annotations is 5246, indicating that about 6.5% of the annotated sentences were annotated with multiple items. Annotation density (annotations per sentence) is 0.48 (1.08, if only the sentences with at least one annotation are considered).

**Table 1:**
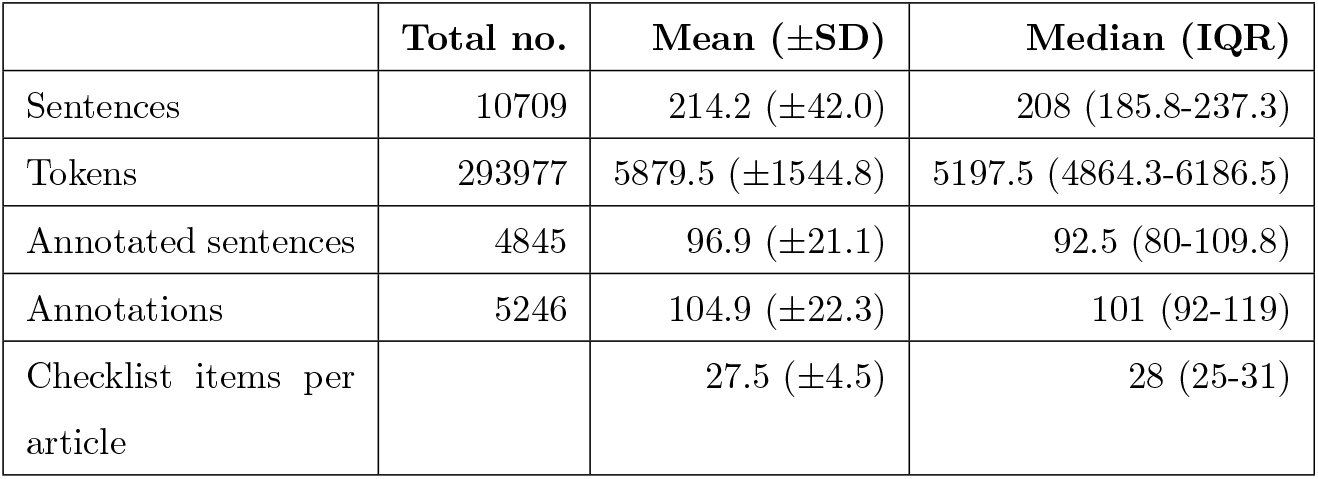
Descriptive statistics regarding 50 manually annotated RCT articles in CONSORT-TM.

The highest number of items associated with a sentence is 5. For instance, the sentence below was labeled with the items Trial Design (3a), Sequence Generation (8a), Allocation Concealment (9), Randomization Implementation (10), and Similarity of Interventions (11b).

1. *Patients were randomly assigned, using a computer-generated randomization schedule, from a central location utilizing an interactive voice response system with blinded medication kit number allocation in a 2:1 ratio to identical-appearing tablets of HZT-501 (800 mg ibuprofen and 26*.*6 mg famotidine) or ibuprofen (800 mg) thrice daily for 24 weeks*.

We found that out of 37 fine-grained items, a median of 28 were annotated per article. The most complete article included 35 CONSORT items, missing discussion relating to Changes to Trial Design (3b) and Interim Analyses/Stopping Guidelines (7b). Since both can be considered contingent items, we considered this article the only one in CONSORT-TM that was fully compliant with the CONSORT guidelines.

Table 2 shows the number of articles annotated with each checklist item and the average number of sentences per article annotated for the item. The results show that 80% of the articles identified themselves as RCTs in the title (n=40) and almost all had structured abstracts (n=49). The major PICO characteristics captured as CONSORT checklist items (Eligibility Criteria (4a), Interventions (5), and Outcomes (6a)) were well-reported (n=49, n=50, n=50, respectively). Similarly, all articles discussed Trial Design (3a) and Statistical Methods for Outcome Comparison (12a) and reported Baseline Data (15) and Outcome Results (17a). Items that indicate some modification to the trial after its launch were infrequently reported. These include Changes to Trial Design (3b) (n=4), Changes to Outcomes (6b) (n=5), and Trial Stopping (14b) (n=6). Protocol Access (24) was the least reported item (n=7), among the rest. Randomization and masking-related items (8a to 11a), which were often found to be reported inadequately in earlier studies, were moderately reported in our collection (60%-80%), except Allocation Concealment (9) (n=19, 38%).

**Table 2:**
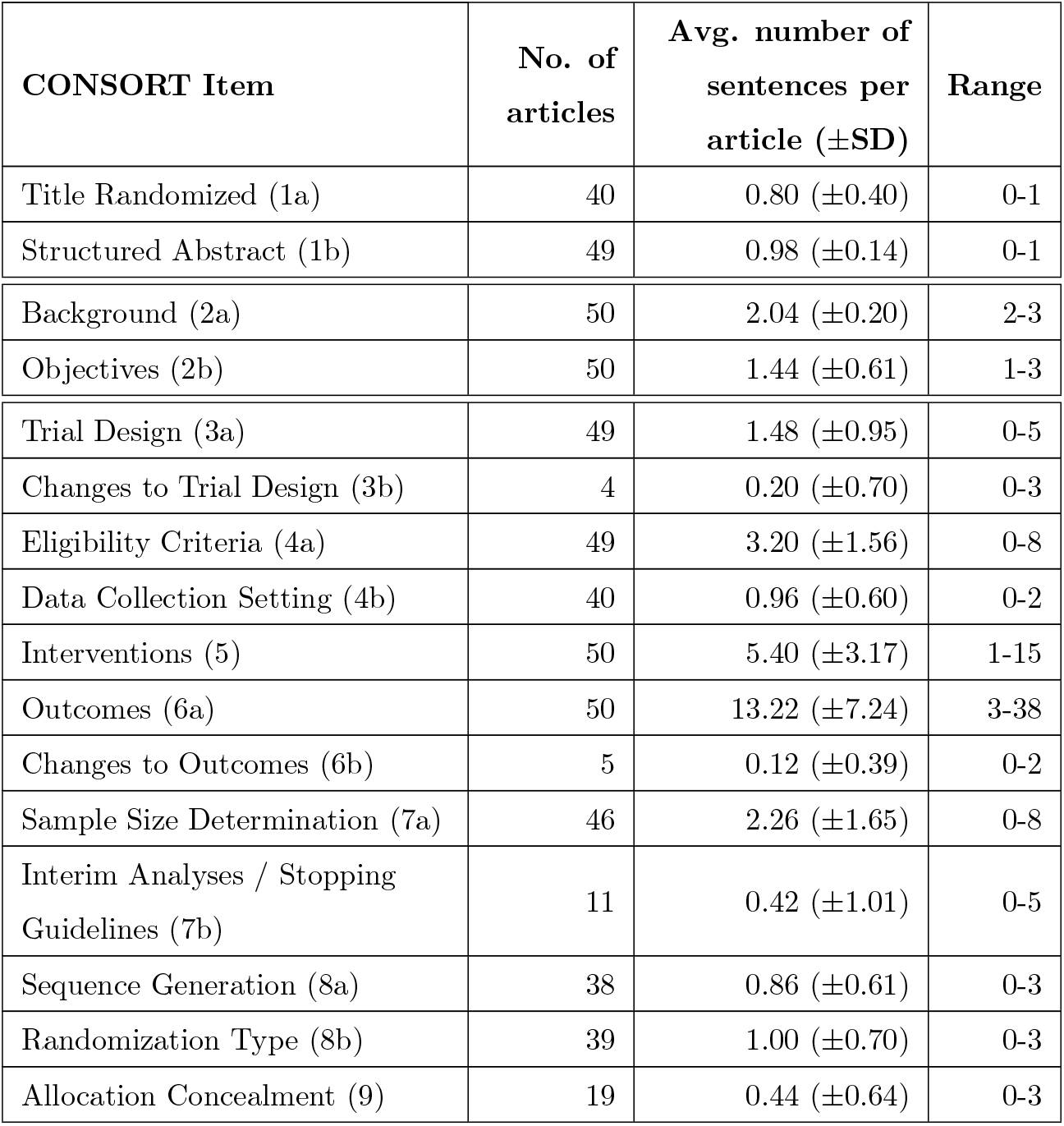
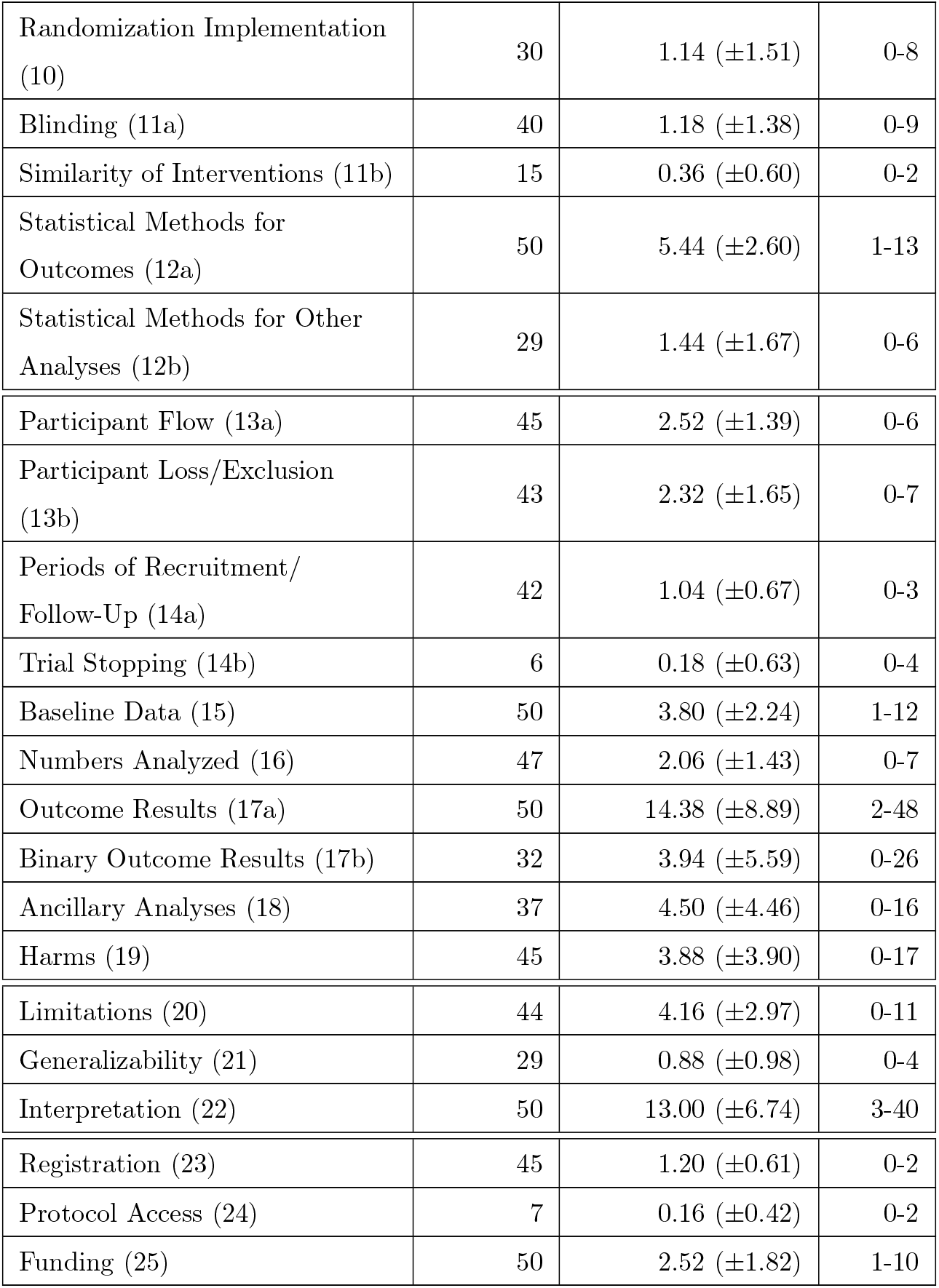
Descriptive statistics regarding the annotation of CONSORT checklist items in CONSORT-TM. SD: standard deviation.

When annotated at all, a checklist item is labeled over 3.47 sentences on average per article (range: 1.13-14.38). While most are annotated in a few sentences (median: 2.19, IQR: 1.33-4.31), a few items are annotated in more than 10 sentences per article (Outcomes (6a), Outcome Results (17a), and Interpretation (22)). We find that items annotated over many sentences cover several aspects of the RCT or have been defined somewhat broadly in CONSORT. For example, the Outcomes category (6a) covers not only primary and secondary outcome measures, but also how and when they were assessed, leading to a large number of sentences being labeled with this item. Similarly, Interpretation (22) is defined as “interpretation consistent with results, balancing benefits and harms, and considering other relevant evidence,” which can encompass the majority of sentences in Discussion sections.

Annotators were instructed to label CONSORT items in the sections with which they are typically associated (excluding those in Other category in CONSORT, such as Registration (23), which could be annotated in any section). However, CONSORT items were not always reported in their associated sections by the authors. This most commonly occurred with Periods of Recruitment and Follow-up (14a) (n=28), Numbers Analyzed (16) (n=15), Participant Flow (13a) (n=12), and Data Collection Setting (4b) (n=11). The first three were often reported in Methods sections, instead of the Results sections, and the latter vice versa.

### Inter-annotator agreement

Inter-annotator agreement at the article level was moderate (mean MASI= 0.60, median=0.63) (Table 3). An interim analysis after 10 articles were double-annotated and adjudicated showed a median MASI of 0.57, indicating that inter-annotator agreement improved over the course of the study. Agreement was highest in back matter sections (Acknowledgements, etc.), where Registration (23) or Funding (25) were often reported (section mean MASI=0.89). While the agreement in Introduction sections was low in the interim analysis (mean MASI=0.50), it improved after guidelines about annotating Background (2a) were clarified (MASI=0.67). Agreement in Methods and Discussion sections were similar (mean MASI of 0.59 and 0.58, respectively), with more spread in agreement in Discussion sections. The agreement in Results sections was lowest (mean MASI=0.50). The agreement of each annotator with other annotators was largely similar (mean annotator MASI=0.60, range: 0.58-0.64). The annotator who annotated a single article was excluded from this calculation.

**Table 3:**
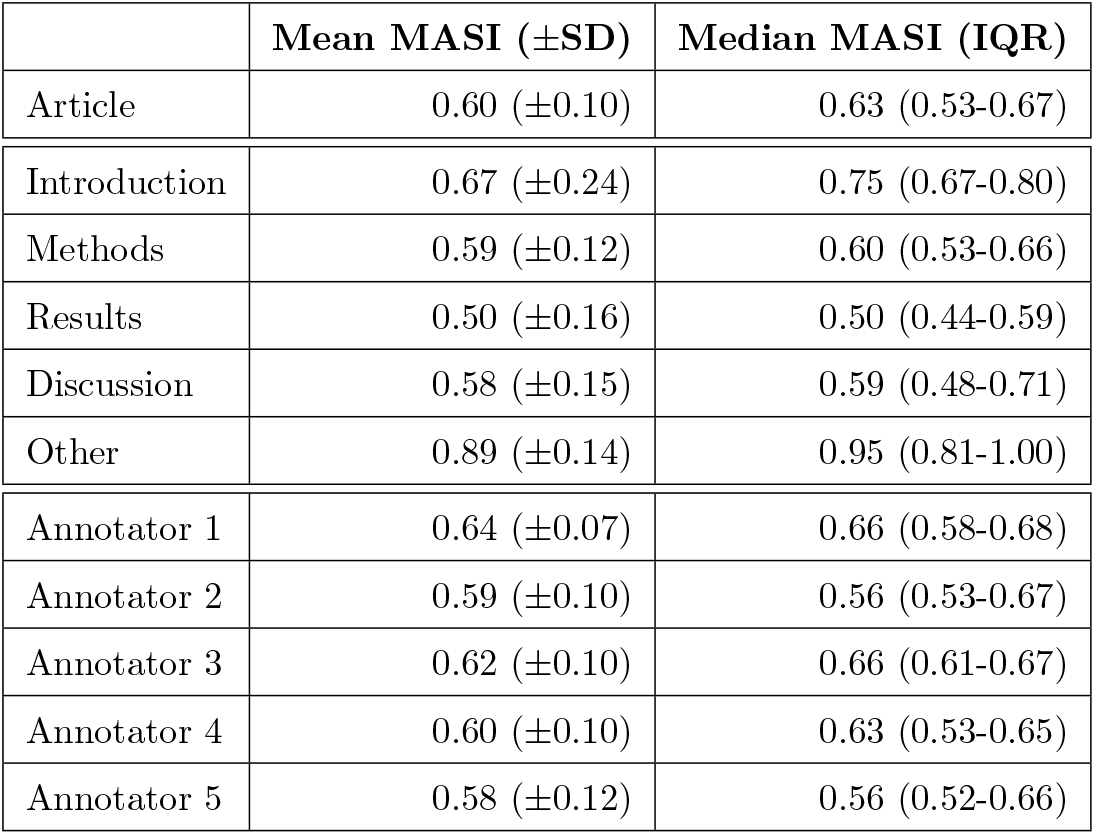
Inter-annotator agreement calculated by MASI formulation. SD: standard deviation; IQR: inter-quartile range.

At the checklist item level (Figure 2), agreement was highest for Registration (23) (Krippendorff’s *α*=0.96). There was also high agreement for Sample Size Determination (7a) (*α*=0.85), followed by Eligibility Criteria (4a) (*α*=0.79) and Objectives (2b) (*α*=0.71). Agreement was lowest for Numbers Analyzed (16) (*α*=0.06), Generalizability (21) (*α*=0.14), and Binary Outcome Results (17b) (*α*=0.15). Average *α* was 0.47.

**Figure 2:**
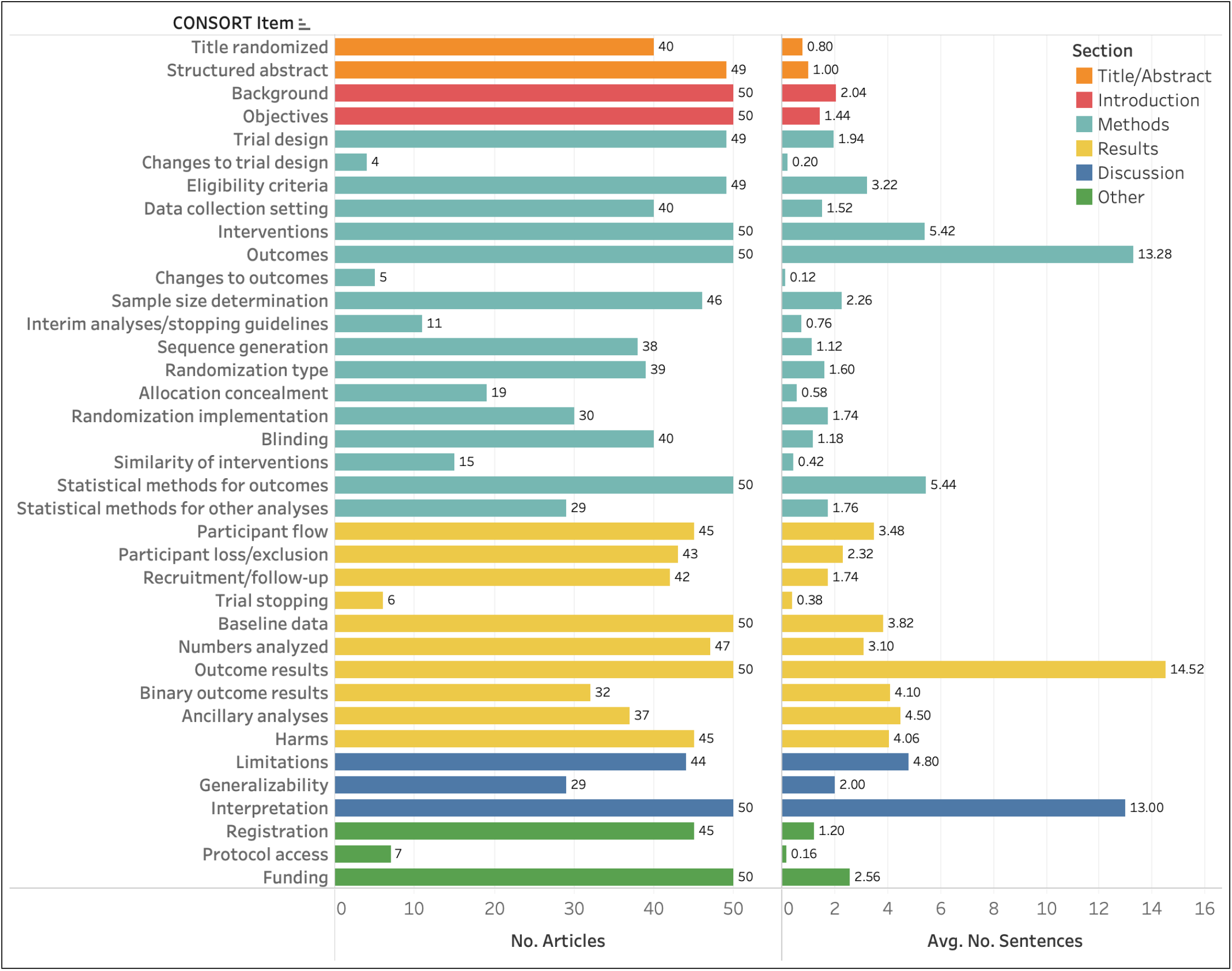
Inter-annotator agreement at the CONSORT item level, calculated using Krippendorff’s *α*.Items are color coded by their associated sections, as sown in the legend.

### Text mining results

The results of baseline experiments with four methods are shown in Table 4. The phrase-based method largely outperformed the section header-based method, especially for rare items (e.g., Changes to Outcomes), which are unlikely to be discussed in dedicated subsections. In contrast, the section header-based method performed better for common items that are often discussed in dedicated subsections (Outcomes). Both of these methods were mostly outperformed by the supervised learning algorithms, with the BioBERT-based model performing best overall in all categories (0.82 precision, 0.63 recall, 0.72 F_1_ score, 0.812 AUC). This model performed particularly well for items with larger number of annotations (Interventions, Outcomes, Statistical Methods for Outcomes). However, it performed poorly for rare items yielding no correct predictions for several items, which led to its lower macro-averaged performance, compared to macro-averaged performance of the phrase-based method and the linear SVM classifier. On the whole, we obtained best performance for Sample Size Determination. Model performance differences on individual items (calculated using McNemar’s test) were largely statistically significant at 95% confidence level, excluding the infrequent items, as shown in Table 4.

**Table 4:**
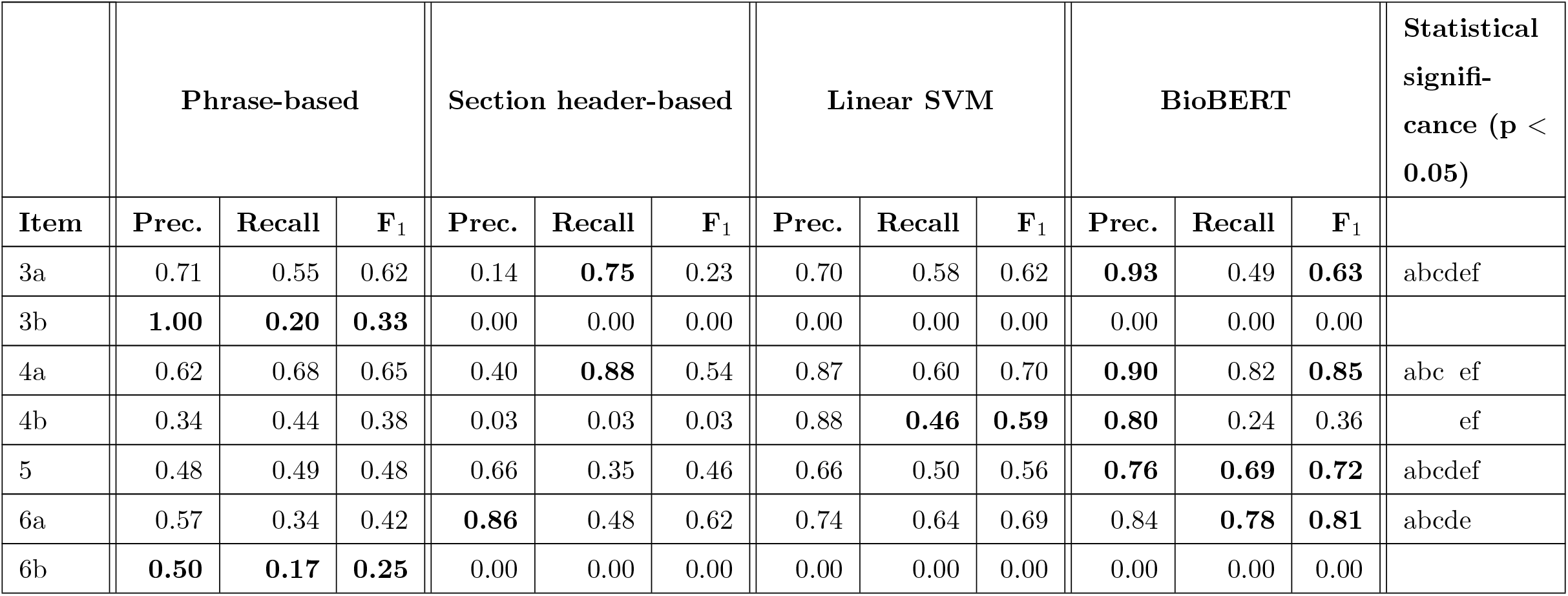
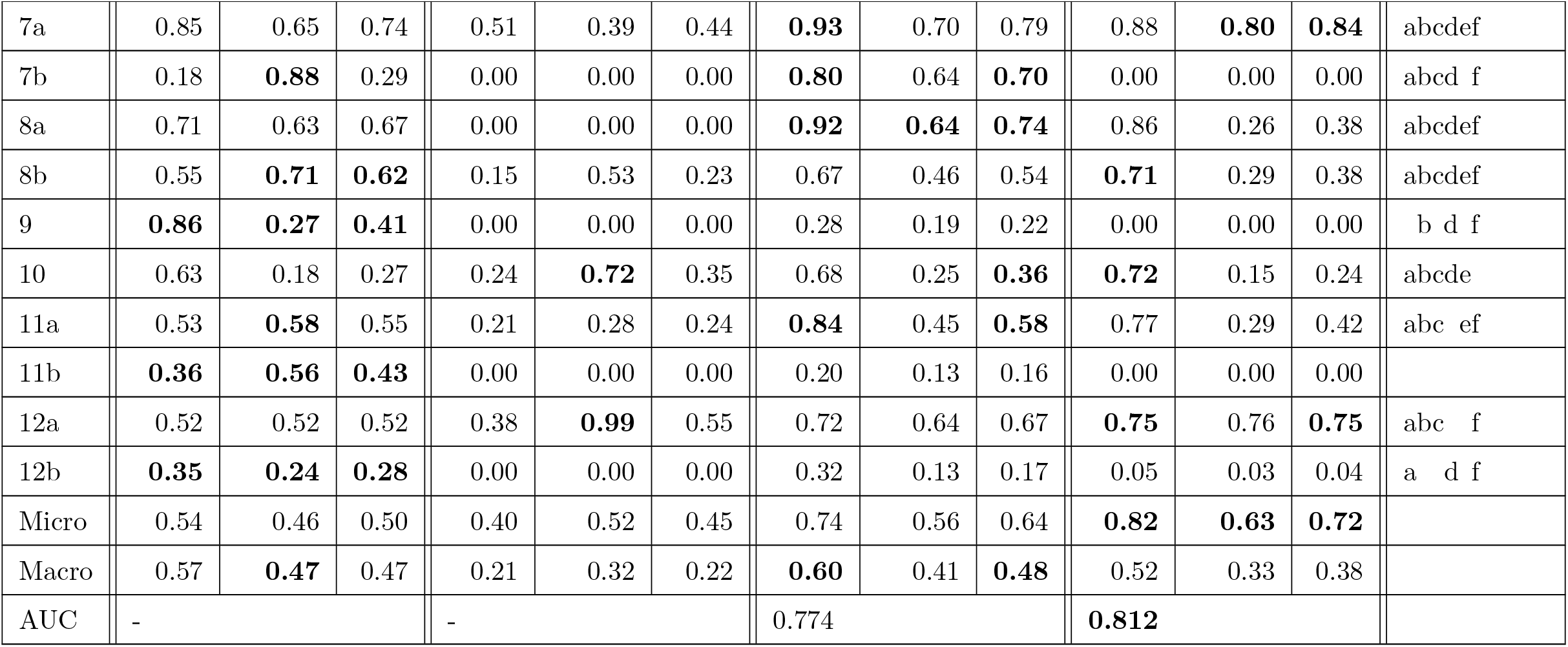
Baseline experiment results. The results for phrase-based and section header-based methods were obtained from all 50 articles, and the results for linear SVM and BioBERT-based models were obtained using 5-fold cross-validation. Best results for each CONSORT item are in bold. 3a: Trial Design; 3b: Changes to Trial Design; 4a: Eligibility Criteria; 4b: Data Collection Setting; 5: Interventions; 6a: Outcomes; 6b: Changes to Outcomes; 7a: Sample Size Determination; 7b: Interim Analyses/Stopping Guidelines; 8a: Sequence Generation; 8b: Randomization Type; 9: Allocation Concealment; 10: Randomization Implementation; 11a: Blinding Procedure; 11b: Similarity of Interventions; 12a: Statistical Methods for Outcomes; 12b: Statistical Methods for Other Analyses; Micro: Micro-averaging; Macro: Macro-averaging. AUC: Area Under Receiver Operator Characteristic (ROC) Curve. In the last column, each letter indicates that the results of one method is statistically significantly different from those of another method at 95% confidence level, as measured by McNemar’s test (a: phrase-based vs. section header-based; b: phrase-based vs. linear SVM; c: phrase-based vs. BioBERT; d: section-header based vs. linear SVM; e: section-header based vs. BioBERT; f: linear SVM vs. BioBERT).

The results based on model combinations are provided in Table 5. We only provide the micro- and macro-averaged results for these combinations. Majority vote yields the best precision overall (0.78 micro, 0.74 macro) among combination methods, while the aggregation of all method predictions yields the best recall (0.87 micro, 0.71 macro). Linear SVM and BioBERT model combination (SVM+BERT) improves upon the best base model by about 2 F_1_ points (0.72 to 0.74).

**Table 5:** Results of combining four base models. The results for the base models are also provided for comparison. Best base model performances as well as best combination performances are in bold. PHR: phrase-based method; SECT: section header-based method; SVM: linear SVM model; BERT: BioBERT-based model.

| Combination | Micro-averaging |  |  | Macro-averaging |  |  |
| --- | --- | --- | --- | --- | --- | --- |
| | Prec. | Recall | $F_1$ | Prec. | Recall | $F_1$ |
| PHR | 0.54 | 0.46 | 0.50 | 0.57 | **0.47** | 0.47 |
| SECT | 0.40 | 0.52 | 0.45 | 0.21 | 0.32 | 0.22 |
| SVM | 0.74 | 0.56 | 0.64 | **0.60** | 0.41 | **0.48** |
| BERT | **0.82** | **0.63** | **0.72** | 0.52 | 0.33 | 0.38 |
| Majority Vote | **0.78** | 0.68 | 0.73 | **0.74** | 0.46 | 0.52 |
| PHR+SEC | 0.40 | 0.72 | 0.51 | 0.45 | 0.63 | 0.43 |
| PHR+SVM | 0.57 | 0.71 | 0.63 | 0.57 | 0.60 | 0.54 |
| PHR+BERT | 0.61 | 0.76 | 0.68 | 0.59 | 0.59 | 0.54 |
| SEC+SVM | 0.45 | 0.74 | 0.56 | 0.46 | 0.54 | 0.42 |
| SEC+BERT | 0.47 | 0.75 | 0.58 | 0.30 | 0.45 | 0.31 |
| SVM+BERT | 0.73 | 0.74 | **0.74** | 0.67 | 0.50 | 0.54 |
| PHR+SEC+SVM | 0.41 | 0.82 | 0.55 | 0.44 | 0.69 | 0.46 |
| PHR+SEC+BERT | 0.42 | 0.84 | 0.56 | 0.45 | 0.67 | 0.45 |
| PHR+SVM+BERT | 0.58 | 0.81 | 0.68 | 0.57 | 0.65 | **0.56** |
| SEC+SVM+BERT | 0.46 | 0.82 | 0.59 | 0.47 | 0.58 | 0.44 |
| PHR+SEC+SVM+BERT | 0.42 | **0.87** | 0.56 | 0.44 | **0.71** | 0.47 |

## Discussion

### Annotation

We obtained moderate agreement in annotating fine-grained CONSORT checklist items on average. However, agreement varied considerably between different items. It was tempting to annotate at a more coarse-grained level to achieve higher agreement. We did not do this, as our primary goal was to evaluate CONSORT in its entirety as annotation target and ultimately we would like to address all aspects of RCT reporting. This clearly made the annotation task more difficult and reduced agreement.

We had highest agreement on well-defined items, such as Objectives (2b), Sample Size Determination (7a), and Registration (23). Agreement was lowest on Numbers Analyzed (16), often only reported in tables/figures. While the annotators were instructed to annotate the captions in these cases, this was not done consistently. Some categories with low agreement are those that are easy to confuse with others. For example, it is challenging to determine whether an outcome result sentence should be labeled as Outcome Result (17a), Binary Outcome Result (17b), or Ancillary Analyses (18) (*α*’s of 0.41, 0.15, and 0.23), as this requires keeping track of all different outcomes and analyses discussed in an article while annotating, a cognitively demanding task. We found that it was easy to over-annotate some broadly conceived items (Background, Interventions, Outcomes, Interpretation). We instructed annotators to limit Background annotations to the two most representative sentences as these sentences generally did not directly relate to the RCT study under consideration. We did not do so for other items to avoid missing important study characteristics, although it may be reasonable to limit the annotation of broadly conceived items to the most representative sentences.

Low agreement on some items may raise questions about the reliability of the annotations and their use for text mining, as *α* coefficient lower than 0.67 is sometimes considered unreliable. We note that the sentences on which agreement was measured were further adjudicated prior to being used for text mining and the rest were examined by two annotators; therefore, we expect that the input labels for text mining methods should be reasonably reliable. Of course, it would be ideal for all annotators to discuss all disagreements to reach a consensus; however, given the complexity of the task, this was not deemed feasible in this study. It is also worth noting that *α*, originally developed for content analysis, emphasizes replicability of annotations, whereas the focus is generally on usefulness in developing natural language processing corpora [48]. Finally, reported inter-annotator agreement in biomedical corpora focusing on similar phenomena is comparable to ours. For example, in the PIBOSO-NICTA corpus [31], Cohen’s *κ* was measured as 0.71, 0.63, 0.61, and 0.41 for Outcomes, Population, Intervention, and Study Design, respectively. We calculated *κ* for the corresponding CONSORT items (6a, 4a, 5, 3a) and obtained 0.56, 0.82, 0.53, and 0.64, respectively. Note also that their annotation focused on abstracts only, which are arguably much easier to annotate than full-text articles.

Nevertheless, low agreement on some items suggests that using CONSORT checklist items as annotation target as-is may not be the most appropriate strategy. For alternative categorizations, we investigated whether higher inter-annotator agreement could be achieved by collapsing related items into a single category. Higher agreement was achieved for the following combinations:

- Randomization Type (8b) and Randomization Implementation (10): combined *α*=0.50 vs. 0.48 and 0.35, respectively.
- Statistical Methods for Outcomes (12a) and Statistical Methods for Other Analyses (12b): combined *α*=0.67 vs. 0.53 and 0.28, respectively.
- Participant Flow (13a) and Participant Loss/Exclusion (13b): combined *α*= 0.51 vs. 0.44 and 0.45, respectively.
- Outcome Result (17a), Binary Outcome Result (17b), and Ancillary Analyses (18): combined *α*= 0.56 vs. 0.41, 0.15, and 0.23, respectively.

From an annotation and text mining perspective, it may be beneficial to combine these categories, as this would lead to more consistent annotations and likely better text mining performance; however, this needs to be weighed against whether the resulting categorization would still serve the use cases under consideration.

On a related note, we also considered merging several infrequent and contingent items into their more frequent siblings. These combinations were:

- Changes to Trial Design (3b) into Trial Design (3a)
- Changes to Outcomes (6b) into Outcomes (6a)
- Trial Stopping (14b) into Recruitment/Follow-Up (14a)

This did not affect inter-annotator agreement significantly, since there were only a small number of annotations for the first items. However, from a text mining standpoint, it may be also be advantageous to merge these categories.

We also noted that some important trial characteristics are not captured by CONSORT guidelines. One example is the lack of a checklist item corresponding to research ethics and consent statements, which are often reported in publications. While it is pointed out in the CONSORT explanation and elaboration article [19] that this is by design, we believe it would be important to include this as an additional category.

While we did not formally measure the time it takes to annotate an article, we found anecdotally that it took about 2-3 hours of concentrated effort. Some variation is to be expected based on annotator’s knowledge of the subject matter discussed in the article and clinical trial methodology. Despite the annotation challenges, we believe that CONSORT-TM can be useful as a benchmark for automated systems that aim to measure CONSORT adherence or extract study characteristics of RCTs. Considering that it involves 37 categories, CONSORTTM can also serve as a challenging biomedical sentence classification corpus.

### Text mining methods

Supervised classification methods had a clear advantage for common checklist items (e.g., Interventions, Outcomes), while only the phrase-based method performed relatively well for infrequent items. This confirms the need for large quantities of labeled data for training effective supervised learning models. We found that approaches that combine predictions from different base models can also improve classification performance. An effective approach could be to rely on the phrase-based method for predicting rare items and combine those with predictions from the BioBERT-based model.

We used standard settings for supervised learning, and it may be possible to achieve better performance with more advanced features or modeling approaches. In the case of SVM classification, we experimented with semantic features derived from MetaMap [49] (entities and their semantic types extracted from sentences). Semantic types features slightly improved results (although not statistically significantly; results not shown), whereas concept features caused a minor degradation. In another approach, we can cast the problem as a sequence labeling task, leveraging the fact that discussion of items often follows a predictable sequence (e.g., Methods sections generally begin with Study Design sentences).

In training the BioBERT-based model, a simple sigmoid layer was used on top of the BERT encoder for classification, which can be substituted by more layers or a more complex neural architecture, such as convolutional or recurrent neural network (CNN or RNN) for higher classification performance. Note, however, that the BioBERT model is already much more complex than the other methods reported here, and takes orders of magnitude longer to train compared to the SVM classifier (hours vs. seconds). Improvements due to additional layers or architectural features may not be sufficiently large to justify the added complexity.

Overall, our preliminary results were encouraging, although it is clear that there remains much room for improvement. Performance for several items may be acceptable for practical use (e.g., Eligibility Criteria, Sample Size Determination), whereas more work is needed for others.

A promising direction may be to use weak supervision to automatically annotate a large number of clinical trial publications using simple heuristics, such as the phrase-based or section header-based methods and then use the resulting (somewhat noisy) data to train more effective classifiers. We have shown that this improves the performance of recognizing sample size and power calculation statements [50], and we plan to extend it to recognition of other CONSORT items as well. Data augmentation [51] can also be used to generate a larger set of (synthetic) examples for infrequent items, which may also benefit machine learning performance.

### Limitations

Our study has several limitations. First, our dataset consists of a small number of articles from 11 journals, which may not be representative of all RCT articles. While we collected more than 5K CONSORT annotations from this set, making this a moderate-sized corpus amenable to data-driven approaches, we acknowledge that text mining approaches would benefit from a larger corpus, considering the large number of categories. Second, our approach focuses solely on textual elements. Much important information about RCTs is in tables, figures, and increasingly in supplementary material. We attempted to address the former two by caption annotation, but we have not tried to incorporate supplementary material or other trial resources. Third, we only annotated whether a sentence discussed a particular checklist item. We did not annotate whether the item is adequately discussed or whether the study fulfills rigor criterion for that item. For instance, a sentence indicating that no blinding was performed was still annotated with the item Blinding Procedure (11a). While this would work well in a human-in-the-loop system where an expert assesses rigor based on sentences highlighted by our methods, it cannot automatically determine whether the study is sufficiently rigorous. Finally, as discussed, our classification methods were preliminary and, for the most part, their accuracy needs to be further improved for practical use.

## Conclusion

We presented CONSORT-TM, a corpus annotated with CONSORT checklist items, and studied baseline sentence classification methods as well as their combinations to recognize a subset of these items. By adopting all the checklist items as our annotation target, we created a corpus with more granular clinical trial information compared to earlier similar efforts [28, 29, 31, 35], one which can facilitate more comprehensive analysis of rigor and transparency of RCT articles. Because CONSORT has been developed and refined by a consortium of trialists, methodologists, and journal editors and reflects their recommendations and practical information needs, we believe that CONSORT-TM can support automated approaches to RCT assessment as well as authoring assistance. On the other hand, our experience showed some challenges in annotating against CONSORT, which leads us to suggest some refinements that may inform followup studies in using reporting guidelines for annotation and text mining.

In future work, we plan to extend our methods to section other than Methods and explore more sophisticated modeling approaches. Another direction is to extract relevant mention-level information and mapping it to standardized terms/identifiers in controlled vocabularies, such as the Ontology for Biomedical Investigations (OBI) [52] (e.g., instead of labeling a sentence as a Study Design sentence, extracting *double-blind study* as the study design). Such mapping could enable large-scale interrogation of clinical trial reports and reveal long-term trends. The principles learned with CONSORT can also be applied to annotating corpora targeting other reporting guidelines, such as ARRIVE for pre-clinical animal studies [11].

## Data Availability

CONSORT-TM is publicly available at https://github.com/kilicogluh/CONSORT-TM.

## Acknowledgments

We thank Tony Tse for his contribution to guideline development and annotation.

## Footnotes

1 http://www.consort-statement.org/about-consort/endorsers, retrieved on 11/02/2020.

2 In this paper, we capitalize CONSORT checklist item names and indicate the corresponding item number in CONSORT guidelines in parentheses (e.g., Trial Design (3a))

3 https://work.cochrane.org/pubmed

4 https://scikit-learn.org/stable

5 https://github.com/ThilinaRajapakse/simpletransformers

