## Supplementary file 1 for "Toward Assessing Clinical Trial Publications for Reporting Transparency"

### CONSORT-TM Articles by Journal

| | Number of articles | CONSORT endorsement |
|---|---|---|
| American Journal of Gastroenterology | 3 | Yes |
| Annals of the Rheumatic Diseases | 9 | Yes |
| Annals of Surgery | 1 | Yes |
| Archives of Disease in Childhood | 1 | No |
| The BMJ | 12 | Yes |
| Diabetes | 2 | No |
| Heart | 4 | Yes |
| Journal of the American College of Cardiology | 2 | Yes |
| Journal of Neurology, Neurosurgery, and Psychiatry | 3 | Yes |
| The Lancet | 8 | Yes |
| Respiratory Research | 5 | Yes |
| TOTAL | 50 | Yes (9), No (2) |

### CONSORT Checklist Items

The CONSORT checklist items are provided below in Table 1. An example passage corresponding to each item is also given, after the table.

Table 1. CONSORT checklist items, corresponding item numbers, and the sections to which they typically belong.

| Section | Item | Item No | Description |
|---|---|---|---|
| Title | | 1a | Identification as a randomized trial in the title |
| Abstract | | 1b | Structured summary of trial design, methods, results, and conclusions |
| Introduction | Background | 2a | Scientific background and explanation of rationale |
| | Objectives | 2b | Specific objectives or hypotheses |
| Methods | Trial design | 3a | Description of trial design (such as parallel, factorial) including allocation ratio |
| | | 3b | Important changes to methods after trial commencement (such as eligibility criteria), with reasons |
| | Participants | 4a | Eligibility criteria for participants |
| | | 4b | Settings and locations where the data were collected |
| | Interventions | 5 | Interventions for each group with sufficient details to allow replication, including how and when they were administered |

| | | | |
|---|---|---|---|
| | Outcomes | 6a | Completely defined pre-specified primary and secondary outcome measures, including how and when they were assessed |
| | | 6b | Any changes to trial outcomes after the trial commenced, with reasons |
| | Sample size | 7a | How sample size was determined |
| | | 7b | When applicable, explanation of any interim analyses and stopping guidelines |
| | Randomization: Sequence generation | 8a | Method used to generate the random allocation sequence |
| | | 8b | Type of randomization: details of any restriction (such as blocking and block size) |
| | Randomization: Allocation concealment | 9 | Mechanism used to implement the random allocation sequence (such as sequentially numbered containers), describing any steps taken to conceal the sequence until interventions were assigned |
| | Randomization: Implementation | 10 | Who generated the random allocation sequence, who enrolled participants, and who assigned participants to interventions |
| | Blinding | 11a | If done, who was blinded after assignment to interventions (for example, participants, care providers, those assessing outcomes) and how |
| | | 11b | If relevant, description of the similarity of interventions |
| | Statistical methods | 12a | Methods used to compare groups for primary and secondary outcomes |
| | | 12b | Methods for additional analyses, such as subgroup analyses and adjusted analyses |
| Results | Participant flow | 13a | For each group, the number of participants who were randomly assigned, received intended treatment, and were analyzed for the primary outcome |
| | | 13b | For each group, losses and exclusions after randomization, together with reasons |
| | Recruitment | 14a | Dates defining the periods of recruitment and follow-up |
| | | 14b | Why the trial ended or was stopped |
| | Baseline data | 15 | A table showing baseline demographic and clinical characteristics for each group |
| | Numbers analyzed | 16 | For each group, number of participants included in each analysis and whether the analysis was by original assigned groups |
| | Outcomes and estimation | 17a | For each primary and secondary outcome, results for each group, and the estimated effect size and its precision (such as 95% confidence interval) |
| | | 17b | For binary outcomes, presentation of both absolute and relative effect sizes |

| | | | |
|---|---|---|---|
| | Ancillary analyses | 18 | Results of any other analyses performed, including subgroup analyses and adjusted analyses, distinguishing pre-specified from exploratory |
| | Harms | 19 | All important harms and unintended effects in each group |
| Discussion | Limitations | 20 | Trial limitations, addressing sources of potential bias, imprecision, and if relevant, multiplicity of analyses |
| | Generalizability | 21 | Generalizability (external validity, applicability) of the trial findings |
| | Interpretation | 22 | Interpretation consistent with results, balancing benefits and harms, and considering other relevant evidence |
| Other | Registration | 23 | Registration number and name of trial registry |
| | Protocol | 24 | Where the full trial protocol can be accessed, if available |
| | Funding | 25 | Sources of funding and other support (such as supply of drugs), role of funders |

**CONSORT-TM Annotation Guidelines**

*Halil Kilicoglu*

In this project, we annotate a corpus of randomized controlled trial articles (RCTs) to be used in training and evaluation of text mining tools that can automatically identify whether reporting guidelines have been followed in the article. We focus as our target on the CONSORT statement for RCTs which includes a 25-item checklist and a flow diagram. The checklist consists of items on methodological details (e.g., eligibility criteria, outcomes, randomization) and interpretation (e.g., limitations, generalizability), among others.

The annotation is at the sentence level. We assign one or more CONSORT checklist item categories to each sentence (or none). We use an in-house, sentence-level annotation tool that has been updated based on earlier comments (accessible at https://skr1.nlm.nih.gov/ConsortAnnotation).

In this study, each annotator is assigned 10 full-text articles of intervention studies published in several journals, such as BMJ, Lancet, and Annals of the Rheumatic Diseases[1]. Each article will be annotated by two annotators, and each annotation pair will annotate at least one article in common. The annotator should select their name from the drop-down list to start annotating. By using the 'Test' user, annotators can familiarize themselves with the tool and the annotations.

[1] Selected based on their CONSORT endorsement and availability of their articles in XML in PubMed Central.

An article to annotate can be selected from the list at the top. IDs of the articles with some annotations on them are shown in green in the list. Each article is presented on several tabs corresponding to the article sections (Title, Abstract, Introduction, Methods, etc.). The annotator is expected to label each sentence with the appropriate checklist item(s) presented in the drop-down list next to each sentence. Each checklist item is indicated with a name and ID number (Item and Item N° in Table 1). Before starting with the annotation, familiarize yourself with the checklist items.

Several points are worth making with regard to sentence annotation and the interface:

- If a sentence does not fit neatly into any of the categories, it should not be annotated, that is, no items from the drop-down list should be selected.
- Sentence splitting is done automatically, and there may be some errors[2]. If a sentence relevant for a checklist item is erroneously split into two (or more), annotate both (or all) sub-parts of the actual sentence with the applicable item.
- Subsection titles (‘Study design’, ‘Participants’) should normally not be annotated. We are mainly interested in sentences that have actual content regarding checklist items (i.e., what the study design is exactly, instead of whether the sentence simply has the phrase ‘study design’)
- An item should be annotated in the section it typically belongs to in the CONSORT statement (e.g., Trial design (3a) in Methods). However, if an item is not addressed in the section it typically belongs to but only in another section, it can be annotated (an example may be Recruitment/FollowUp, which often is indicated in the Methods, not in Results).
    - Items in the Other category (Registration, Funding, Protocol) can be annotated in any section.
    - In the abstract, annotate checklist item 1b, if appropriate. In addition, if an item is only mentioned in the abstract (such as trial registry), annotate that as well.
- For convenience, the annotator can type the Item N° (see Table 1) for a given checklist item, while in the category drop-down box, instead of scrolling up/down.
- If information relevant to a checklist item appears in a Table or a Figure, annotate the caption of the Table or Figure, if available. Otherwise, the sentence in the narrative text with reference to the Table or Figure can be annotated.
- Captions can be multiple-sentence, and some sentences may not be relevant, like explanation of symbols used in the table, etc. Avoid annotating these sentences.
- Some articles have summary sections such as ‘What this study adds’, ‘What is already known on this topic’, etc. Unless they address items that do not appear anywhere else, they should not be annotated.
- Some sentences need additional context for interpretation (e.g., ‘*This difference in cure rate could be attributed to different populations recruited to the study*.’) Knowing what expressions like ‘this difference’ (i.e., anaphoric expressions) refer to may help interpretation. However, it is not necessary to label the sentence or sentences that detail ‘this difference’ in such cases. It is sufficient to label the sentence above (to prevent over-annotation).

The title of the article is linked to its PDF for reference. The annotator can click on the ‘Item Check’ button to see the list of all CONSORT items and whether they have been annotated in that article (green: annotated, red: not annotated). This can be helpful in identifying missing items.

[2] Though they should be minimal.

Once the annotation of an article is completed, the annotator should click the Submit button, so that the annotations are stored. Note that unless this button is pressed, the results are not saved on the server. So, it makes sense to Submit often.

Some item-specific instructions are below. If you're having difficulty with some items or sentences, it's best to raise the issue with others. These instructions will be expanded as we go along.

- *1a (Identification as a randomized trial in the title)*: Annotate this item always on the first sentence of the title.
- *1b (Whether the abstract is structured for trial design, methods, results, and conclusions)*: Annotate this item always on the first sentence of the abstract, excluding abstract section headers.
- *2a (Scientific background and explanation of rationale):* Background/rationale narrative is typically long, spanning multiple sentences or even multiple paragraphs. Annotate a maximum of 2 sentences that best explain the rationale[3] (i.e., why was this study undertaken?). Different from other items, we are likely to assess this item at the article level (i.e., does the article describe rationale or not?).
- *Outcome-related items (6a, 6b, 12a, 12b, 17a, 17b):* For a clinical trial, it is important to distinguish primary outcomes from secondary outcomes and other additional measures. So, for annotation, it is important to establish what these outcome measures are. This makes it easier to distinguish checklist items pertaining to primary/secondary outcomes from similar items that apply to additional measures (12a vs. 12b, for example).
- *Binary vs. non-binary outcomes (17a vs. 17b)*: Similarly, it is important to distinguish whether an outcome is binary or not (for example, whether a patient recovered completely or not is a binary outcome, whereas the proportion of patients that recover is not).
- *Limitations (20):* Annotate sentences that report specific limitations (small sample size, etc.). If a sentence simply states that the study has limitations, you don't need to annotate it (as in our previous limitations study).

Examples for each item are given below (from "CONSORT 2010 explanation and elaboration: Updated guidelines for reporting parallel group randomised trials" Moher et al., 2010, BMJ)

For another example, see [http://www.consort-statement.org/Documents/SampleView/5eb5d30c-fc53-4708-b0cd-80ab76fe95a5](http://www.consort-statement.org/Documents/SampleView/5eb5d30c-fc53-4708-b0cd-80ab76fe95a5).

Item 1a: Identification as a randomized trial in the title

*Smoking reduction with oral nicotine inhalers: double blind, randomised clinical trial of efficacy and safety*

Item 2a. Scientific background and explanation of rationale

*Surgery is the treatment of choice for patients with disease stage I and II non-small cell lung cancer (NSCLC) … An NSCLC meta-analysis combined the results from eight randomised trials of surgery versus surgery plus adjuvant cisplatin-based chemotherapy and showed a small, but not significant (p=0.08), absolute survival benefit of around 5% at 5 years (from 50% to 55%). At the time the current trial was designed (mid-1990s), adjuvant chemotherapy had not become standard clinical practice … The clinical rationale for neo-adjuvant chemotherapy is three-fold: regression of the primary cancer could be achieved thereby facilitating and simplifying or reducing subsequent surgery; undetected micro-metastases could be dealt with at the start of treatment; and there might be inhibition of the putative stimulus to residual cancer by growth factors released by surgery and by subsequent wound healing … The current trial was therefore set up to compare, in patients with resectable NSCLC, surgery alone versus three*

[3] One criterion to select two sentences would be to consider which 2 sentences would best summarize the rationale for the study.

*cycles of platinum-based chemotherapy followed by surgery in terms of overall survival, quality of life, pathological staging, resectability rates, extent of surgery, and time to and site of relapse.*

Item 2b. Specific objectives or hypotheses
*In the current study we tested the hypothesis that a policy of active management of nulliparous labour would: 1. reduce the rate of caesarean section, 2. reduce the rate of prolonged labour; 3. not influence maternal satisfaction with the birth experience.*

Item 3a. Description of trial design (such as parallel, factorial) including allocation ratio
*This was a multicenter, stratified (6 to 11 years and 12 to 17 years of age, with imbalanced randomisation [2:1]), double-blind, placebo-controlled, parallel-group study conducted in the United States (41 sites).*

Item 3b. Important changes to methods after trial commencement (such as eligibility criteria), with reasons
*Patients were randomly assigned to one of six parallel groups, initially in 1:1:1:1:1:1 ratio, to receive either one of five otamixaban … regimens … or an active control of unfractionated heparin … an independent Data Monitoring Committee reviewed unblinded data for patient safety; no interim analyses for efficacy or futility were done. During the trial, this committee recommended that the group receiving the lowest dose of otamixaban (0·035 mg/kg/h) be discontinued because of clinical evidence of inadequate anticoagulation. The protocol was immediately amended in accordance with that recommendation, and participants were subsequently randomly assigned in 2:2:2:2:1 ratio to the remaining otamixaban and control groups, respectively.*

Item 4a. Eligibility criteria for participants
*Eligible participants were all adults aged 18 or over with HIV who met the eligibility criteria for antiretroviral therapy according to the Malawian national HIV treatment guidelines (WHO clinical stage III or IV or any WHO stage with a CD4 count <250/mm3) and who were starting treatment with a BMI <18.5. Exclusion criteria were pregnancy and lactation or participation in another supplementary feeding programme.*

Item 4b. Settings and locations where the data were collected
*The study took place at the antiretroviral therapy clinic of Queen Elizabeth Central Hospital in Blantyre, Malawi, from January 2006 to April 2007. Blantyre is the major commercial city of Malawi, with a population of 1 000 000 and an estimated HIV prevalence of 27% in adults in 2004.*

Item 5. The interventions for each group with sufficient details to allow replication, including how and when they were actually administered
*In POISE, patients received the first dose of the study drug (i.e., oral extended-release metoprolol 100 mg or matching placebo) 2-4 h before surgery. Study drug administration required a heart rate of 50 bpm or more and a systolic blood pressure of 100 mm Hg or greater; these haemodynamics were checked before each administration. If, at any time during the first 6 h after surgery, heart rate was 80 bpm or more and systolic blood pressure was 100 mm Hg or higher, patients received their first postoperative dose (extended-release metoprolol 100 mg or matched placebo) orally. If the study drug was not given during the first 6 h, patients received their first postoperative dose at 6 h after surgery. 12 h after the first postoperative dose, patients started taking oral extended-release metoprolol 200 mg or placebo every day for 30 days. If a patient's heart rate was consistently below 45 bpm or their systolic blood pressure dropped below 100 mm Hg, study drug was withheld until their heart rate or systolic blood pressure recovered; the study drug was then restarted at 100 mg once daily. Patients whose heart rate was consistently 45-49 bpm and systolic blood pressure exceeded 100 mm Hg delayed taking the study drug for 12 h.*

Item 6a. Completely defined pre-specified primary and secondary outcome measures, including how and when they were assessed
*The primary endpoint with respect to efficacy in psoriasis was the proportion of patients achieving a 75% improvement in psoriasis activity from baseline to 12 weeks as measured by the PASI [psoriasis area and severity*

*index] Additional analyses were done on the percentage change in PASI scores and improvement in target psoriasis lesions.*

Item 6b. Any changes to trial outcomes after the trial commenced, with reasons
*The original primary endpoint was all-cause mortality, but, during a masked analysis, the data and safety monitoring board noted that overall mortality was lower than had been predicted and that the study could not be completed with the sample size and power originally planned. The steering committee therefore decided to adopt co-primary endpoints of all-cause mortality (the original primary endpoint), together with all-cause mortality or cardiovascular hospital admissions (the first prespecified secondary endpoint).*

Item 7a. How sample size was determined
*To detect a reduction in PHS (postoperative hospital stay) of 3 days (SD 5 days), which is in agreement with the study of Lobo et al with a two-sided 5% significance level and a power of 80%, a sample size of 50 patients per group was necessary, given an anticipated dropout rate of 10%. To recruit this number of patients a 12-month inclusion period was anticipated.*
Item 7b. When applicable, explanation of any interim analyses and stopping guidelines
*Two interim analyses were performed during the trial. The levels of significance maintained an overall P value of 0.05 and were calculated according to the O'Brien-Fleming stopping boundaries. This final analysis used a Z score of 1.985 with an associated P value of 0.0471.*

Item 8a. Method used to generate the random allocation sequence
*Independent pharmacists dispensed either active or placebo inhalers according to a computer generated randomisation list.*

Item 8b. Type of randomization; details of any restriction (such as blocking and block size)
*Randomization sequence was created using Stata 9.0 (StataCorp, College Station, TX) statistical software and was stratified by center with a 1:1 allocation using random block sizes of 2, 4, and 6.*

Item 9. Mechanism used to implement the random allocation sequence (such as sequentially numbered containers), describing any steps taken to conceal the sequence until interventions were assigned
*The allocation sequence was concealed from the researcher (JR) enrolling and assessing participants in sequentially numbered, opaque, sealed and stapled envelopes. Aluminium foil inside the envelope was used to render the envelope impermeable to intense light. To prevent subversion of the allocation sequence, the name and date of birth of the participant was written on the envelope and a video tape made of the sealed envelope with participant details visible. Carbon paper inside the envelope transferred the information onto the allocation card inside the envelope and a second researcher (CC) later viewed video tapes to ensure envelopes were still sealed when participants' names were written on them. Corresponding envelopes were opened only after the enrolled participants completed all baseline assessments and it was time to allocate the intervention.*

Item 10. Who generated the allocation sequence, who enrolled participants, and who assigned participants to interventions
*Determination of whether a patient would be treated by streptomycin and bed-rest (S case) or by bed-rest alone (C case) was made by reference to a statistical series based on random sampling numbers drawn up for each sex at each centre by Professor Bradford Hill; the details of the series were unknown to any of the investigators or to the co-ordinator … After acceptance of a patient by the panel, and before admission to the streptomycin centre, the appropriate numbered envelope was opened at the central office; the card inside told if the patient was to be an S or a C case, and this information was then given to the medical officer of the centre.*

Item 11a. If done, who was blinded after assignment to interventions (for example, participants, care providers, those assessing outcomes) and how

*Whereas patients and physicians allocated to the intervention group were aware of the allocated arm, outcome assessors and data analysts were kept blinded to the allocation.*

Item 11b. If relevant, description of the similarity of interventions
*Jamieson Laboratories Inc provided 500-mg immediate release niacin in a white, oblong, bisect caplet. We independently confirmed caplet content using high performance liquid chromatography… The placebo was matched to the study drug for taste, color, and size, and contained microcrystalline cellulose, silicon dioxide, dicalcium phosphate, magnesium stearate, and stearic acid.*

Item 12a. Statistical methods used to compare groups for primary and secondary outcomes
*The primary endpoint was change in bodyweight during the 20 weeks of the study in the intention-to-treat population … Secondary efficacy endpoints included change in waist circumference, systolic and diastolic blood pressure, prevalence of metabolic syndrome … We used an analysis of covariance (ANCOVA) for the primary endpoint and for secondary endpoints waist circumference, blood pressure, and patient-reported outcome scores; this was supplemented by a repeated measures analysis. The ANCOVA model included treatment, country, and sex as fixed effects, and bodyweight at randomisation as covariate. We aimed to assess whether data provided evidence of superiority of each liraglutide dose to placebo (primary objective) and to orlistat (secondary objective).*

Item 12b. Methods for additional analyses, such as subgroup analyses and adjusted analyses
*Proportions of patients responding were compared between treatment groups with the Mantel-Haenszel $\chi 2$ test, adjusted for the stratification variable, methotrexate use.*

Item 13. Participant flow (a diagram is strongly recommended)

Item 13a. For each group, the numbers of participants who were randomly assigned, received intended treatment, and were analysed for the primary outcome

Item 13b. For each group, losses and exclusions after randomisation, together with reasons
*There was only one protocol deviation, in a woman in the study group. She had an abnormal pelvic measurement and was scheduled for elective caesarean section. However, the attending obstetrician judged a trial of labour acceptable; caesarean section was done when there was no progress in the first stage of labour.*

Item 14a. Dates defining the periods of recruitment and follow-up
*Age-eligible participants were recruited … from February 1993 to September 1994 … Participants attended clinic visits at the time of randomisation (baseline) and at 6-month intervals for 3 years*

Item 14b. Why the trial ended or was stopped
*In January 2000, problems with vaccine supply necessitated the temporary nationwide replacement of the whole cell component of the combined DPT/Hib vaccine with acellular pertussis vaccine. As this vaccine has a different local reactogenicity profile, we decided to stop the trial early*

Item 15. A table showing baseline demographic and clinical characteristics for each group

Item 16. For each group, number of participants (denominator) included in each analysis and whether the analysis was by original assigned groups
*The primary analysis was intention-to-treat and involved all patients who were randomly assigned*

Item 17a. For each primary and secondary outcome, results for each group, and the estimated effect size and its precision (such as 95% confidence interval)

Item 17b. For binary outcomes, presentation of both absolute and relative effect sizes is recommended

*The risk of oxygen dependence or death was reduced by 16% (95% CI 25% to 7%). The absolute difference was −6.3% (95% CI −9.9% to −2.7%); early administration to an estimated 16 babies would therefore prevent 1 baby dying or being long-term dependent on oxygen" (also see table 7).*

Item 18. Results of any other analyses performed, including subgroup analyses and adjusted analyses, distinguishing pre-specified from exploratory

*On the basis of a study that suggested perioperative β-blocker efficacy might vary across baseline risk, we prespecified our primary subgroup analysis on the basis of the revised cardiac risk index scoring system. We also did prespecified secondary subgroup analyses based on sex, type of surgery, and use of an epidural or spinal anaesthetic. For all subgroup analyses, we used Cox proportional hazard models that incorporated tests for interactions, designated to be significant at p<0.05 … Figure 3 shows the results of our prespecified subgroup analyses and indicates consistency of effects … Our subgroup analyses were underpowered to detect the modest differences in subgroup effects that one might expect to detect if there was a true subgroup effect.*

Item 19. All important harms or unintended effects in each group

*The proportion of patients experiencing any adverse event was similar between the rBPI21 [recombinant bactericidal/permeability-increasing protein] and placebo groups: 168 (88.4%) of 190 and 180 (88.7%) of 203, respectively, and it was lower in patients treated with rBPI21 than in those treated with placebo for 11 of 12 body systems … the proportion of patients experiencing a severe adverse event, as judged by the investigators, was numerically lower in the rBPI21 group than the placebo group: 53 (27.9%) of 190 versus 74 (36.5%) of 203 patients, respectively. There were only three serious adverse events reported as drug-related and they all occurred in the placebo group.*

Item 20. Trial limitations, addressing sources of potential bias, imprecision, and, if relevant, multiplicity of analyses

*The preponderance of male patients (85%) is a limitation of our study … We used bare-metal stents, since drug-eluting stents were not available until late during accrual. Although the latter factor may be perceived as a limitation, published data indicate no benefit (either short-term or long-term) with respect to death and myocardial infarction in patients with stable coronary artery disease who receive drug-eluting stents, as compared with those who receive bare-metal stents*

Item 21. Generalizability (external validity, applicability) of the trial findings

*As the intervention was implemented for both sexes, all ages, all types of sports, and at different levels of sports, the results indicate that the entire range of athletes, from young elite to intermediate and recreational senior athletes, would benefit from using the presented training programme for the prevention of recurrences of ankle sprain. By including non-medically treated and medically treated athletes, we covered a broad spectrum of injury severity. This suggests that the present training programme can be implemented in the treatment of all athletes. Furthermore, as it is reasonable to assume that ankle sprains not related to sports are comparable with those in sports, the programme could benefit the general population."*

Item 22. Interpretation consistent with results, balancing benefits and harms, and considering other relevant evidence

*Studies published before 1990 suggested that prophylactic immunotherapy also reduced nosocomial infections in very-low-birth-weight infants. However, these studies enrolled small numbers of patients; employed varied designs, preparations, and doses; and included diverse study populations. In this large multicenter, randomised controlled trial, the repeated prophylactic administration of intravenous immune globulin failed to reduce the incidence of nosocomial infections significantly in premature infants weighing 501 to 1500 g at birth.*

Item 23. Registration number and name of trial registry

*The trial is registered at ClinicalTrials.gov, number NCT00244842.*

Item 24. Where the full trial protocol can be accessed, if available

*Full details of the trial protocol can be found in the Supplementary Appendix, available with the full text of this article at www.nejm.org.*

Item 25. Sources of funding and other support (such as supply of drugs), role of funders

*Grant support was received for the intervention from Plan International and for the research from the Wellcome Trust and Joint United Nations Programme on HIV/AIDS (UNAIDS). The funders had no role in study design, data collection and analysis, decision to publish, or preparation of the manuscript.*
